# Multidimensional autonomic dynamics are associated with clinical improvement across psychiatric patients

**DOI:** 10.64898/2026.09.15.26363091

**Authors:** Noa Mauda, Kirill Vasilchenko, Noa Danan, Ihab Darawshy, Shir Galin, Saher Sheikh, Reut Naim, Vladimir Zlidennyy, Lidia Izakson, Hanna Keren

## Abstract

Psychiatric symptoms are heterogeneous and change over time, yet clinical assessment relies largely on discrete subjective symptom assessments. We examined whether affective reactivity and autonomic dynamics relate to current clinical state and subsequent symptom change in a transdiagnostic psychiatric sample. Patients were assessed at baseline using an adaptive reward-based mood modulation task (n = 100) and heart rate variability recording (n = 57), with longitudinal clinical data available approximately 4 weeks later (n=34). Patients showed reduced positive affective reactivity and altered linear and nonlinear HRV. Positive affective reactivity was primarily associated with concurrent affective symptoms, whereas LF/HF ratio, Lyapunov exponent and Approximate Entropy were prospectively associated with subsequent symptom improvement. Integrating these HRV features into a multidimensional index yielded a stronger association with symptom change beyond baseline cross-sectional symptom severity, suggesting that this index may provide relevant information about clinical trajectories during treatment.

## Introduction

Psychiatric disorders are heterogeneous and dynamic, posing major challenges for characterization and longitudinal monitoring with existing assessment tools. Clinical assessment relies predominantly on symptom-based evaluations, typically using structured interviews and rating scales based on diagnostic frameworks such as the DSM-5 and ICD-11 (Cevoli et al., 2025; American Psychiatric Association, 2022; World Health Organization, 2019). While these approaches provide a common diagnostic language, they present two key limitations. First, they rely heavily on subjective symptom reports and provide limited objective information; second, they are typically administered at discrete time points, limiting their ability to capture the continuous dynamics and temporal evolution of mental states (Allsopp et al., 2019; Leucht et al., 2024; Spiller et al., 2024). Consequently, there is a growing need for measures that can complement clinical assessment by reflecting current clinical state and prospectively informing about clinical trajectories and change.

A growing body of research suggests that psychiatric disorders can be conceptualized as disturbances in regulatory processes governing emotional, cognitive, and physiological systems (Cludius et al., 2020; Morawetz et al., 2025; Purcell, 2025). Difficulties in emotion regulation are central to the development and maintenance of psychiatric disorders (Sloan et al., 2017). One key aspect of emotional regulatory processes is emotional updating, the ability to adjust emotional states in response to environmental feedback (Rutledge et al., 2014; Eldar et al., 2016; Keren et al., 2021; Nagar et al., 2026). A recent adaptive experimental paradigm enables the measurement, modeling, and perturbation of this process, providing a quantifiable measure of emotional reactivity and regulatory dynamics (Keren et al., 2021; Nagar et al., 2026). In this paradigm, a closed-loop adaptive task manipulates reward prediction errors (RPEs) to induce personalized changes in momentary mood, providing a tractable framework for probing emotional reward-based reactivity (Keren et al., 2021). In the present study, this paradigm was used to quantify individual differences in emotional reactivity as reward contingencies shifted between positive and negative RPEs.

Autonomic physiology provides a continuous window into bodily regulatory dynamics that are increasingly implicated in psychiatric disorders. Heart rate variability (HRV), a non-invasive measure of autonomic regulation, captures beat-to-beat variation in cardiac timing and can be repeatedly assessed using wearable sensors (Carr et al., 2018; Ramesh et al., 2023). Altered HRV has been associated with a broad range of psychiatric conditions, including depression, anxiety disorders, and psychotic disorders (Gullett et al., 2023; Heiss et al., 2021; Mulcahy et al., 2019; L. Wang et al., 2024). Across these disorders, reduced linear HRV measures are commonly interpreted as reflecting diminished parasympathetic regulation and reduced reactivity to environmental changes (Galin et al., 2026; Lenger et al., 2022).

Many previous studies have focused on conventional linear HRV metrics that estimate the magnitude of variability in cardiac signals, including time-domain measures (e.g., RMSSD) that quantify the magnitude of beat-to-beat variability and frequency-domain measures (e.g., LF/HF ratio) that characterize the spectral distribution of cardiac variability (Galin et al., 2026; Galin & Keren, 2024; Shaffer & Ginsberg, 2017). However, physiological systems also exhibit nonlinear dynamics characterized by complex temporal structures, sensitivity to perturbations, and evolving stability over time. These properties can be quantified using less common nonlinear HRV measures (Silva et al., 2017; Stavrakis et al., 2020).

Among these measures, the Lyapunov Exponent (LyapExp) quantifies the sensitivity of a system to small perturbations, with higher values indicating greater divergence of nearby trajectories in the system’s state space (Pham et al., 2025; Valenza et al., 2015; Zhao et al., 2019). Approximate Entropy (ApEn) quantifies the unpredictability of beat-to-beat heart rate fluctuations, with higher values reflecting more unpredictable dynamics. By characterizing temporal organization rather than the magnitude or spectral distribution of variability, ApEn can capture properties of cardiac dynamics not reflected in linear HRV measures. Its sensitivity has been demonstrated, for example, by distinguishing reduced heart rate complexity in infants at risk for sudden infant death syndrome from that of healthy infants (Pincus et al., 1993).

Despite growing interest in emotional and autonomic dynamics in psychiatric research (Houben et al., 2015; Mulcahy et al., 2019; Ramesh et al., 2023; Sperry et al., 2020), and increasing efforts to identify clinically useful biomarkers in psychiatry (Abi-Dargham et al., 2023; García-Gutiérrez et al., 2020), it remains unclear which emotional and autonomic measures provide clinically relevant information for psychiatric assessment and monitoring.

There has been increasing emphasis on dimensional and transdiagnostic approaches to psychopathology, which focus on symptom severity rather than categorical diagnoses alone (Cuthbert & Insel, 2013; Fox et al., 2025; Wise et al., 2023). These questions are particularly relevant during psychiatric hospitalization, a period characterized by substantial symptom change under intensive treatment (Melchior et al., 2016; Nelson et al., 2017; Oh et al., 2020). This setting therefore provides a unique opportunity to evaluate different clinical states and trajectories within a heterogeneous psychiatric population. Consistent with the transdiagnostic perspective, the present study adopted a dimensional framework, examining variation in symptom severity across psychiatric patients in addition to categorical comparisons with matched healthy controls.

Overall, we examined whether these measures could (1) distinguish psychiatric patients from healthy controls, (2) relate to symptom severity, (3) prospectively inform subsequent clinical improvement during hospitalization, and (4) show reliability across repeated assessments.

## Methods

### Participants

Participants included a transdiagnostic sample of adult psychiatric patients receiving treatment in inpatient and day-clinic psychiatric services at Ziv medical centre in northern Israel, together with a matched healthy control group recruited from the community (see Table 1 for participants characteristics). Inclusion criteria were age 18-65 years, capacity to provide informed consent, and ability to complete the experimental task. Participants with severe cognitive impairment preventing task completion were excluded. Healthy controls reported no current psychiatric diagnosis and no regular use of psychotropic medication.

**Table 1.** Demographic characteristics of the cohort and the different analysis samples.

| Characteristic | Baseline<br>behavioral<br>sample | Baseline<br>physiological<br>sample | Prospective<br>sample | Healthy<br>controls |
| --- | --- | --- | --- | --- |
| **n** | 100 (of 103<br>recruited) | 57 (of 100) | 34 (of 57) | 29 (of 30<br>recruited) |
| **Age Mean ± SD** | 36.79±12.36 | 35.79±11.92 | 35.60±11.72 | 36.69±11.36 |
| **Female n (%)** | 53 (53%) | 31 (54.4%) | 20 (58.8%) | 20 (69%) |

A total of 103 psychiatric patients were recruited. Of these, 100 completed the behavioural task at baseline and constituted the baseline behavioral analysis sample (53 females, 47 males; mean age = 36.79 ± 12.36 years). The sample represented a broad transdiagnostic cohort comprising depressive disorders (n = 29), schizophrenia spectrum disorders (n = 26), post-traumatic stress disorder (PTSD; n = 15), bipolar disorder (n = 10), personality disorders (n = 10), obsessive-compulsive disorder (OCD; n = 7), and other psychiatric diagnoses (n = 3). In accordance with the dimensional framework of the study, psychiatric diagnoses were used to characterize the sample but were not employed as grouping variables in the statistical analyses.

Of the baseline patient sample, 57 participants had valid baseline HRV recordings (31 females, 26 males; mean age = 35.79 ± 11.92 years) and comprised the baseline physiological analysis sample. Prospective analyses were conducted in the 34 participants with both valid baseline HRV recordings and complete clinical assessments at follow-up. Test–retest reliability analyses included 31 participants, as three individuals from the prospective sample did not have usable HRV recordings at the second assessment.

Healthy control participants were recruited from the community through advertisements and matched to the patient sample on age and sex. Twenty-seven healthy controls completed the behavioural task (18 females, 9 males; mean age = 36.15 ± 11.83 years), and 29 had valid HRV recordings (20 females, 9 males; mean age = 36.69 ± 11.36 years).

All participants provided written informed consent prior to participation. The study was approved by the local institutional ethics committee (approval no. Ziv-0068-23).

### Study design and procedure

Patients completed the experimental protocol at baseline and again at follow-up during hospitalization approximately one month later (mean interval = 27.16 ± 9.43 days), allowing assessment of clinical change over the course of treatment. Healthy controls completed a single experimental session under identical experimental conditions (Fig. 1A). At each session, participants completed the behavioral task (Fig. 1B) while physiological signals were recorded continuously throughout the experiment. Clinical symptom severity was assessed by trained clinicians on the same day whenever possible, or within a maximum interval of two days.

**Figure 1.**
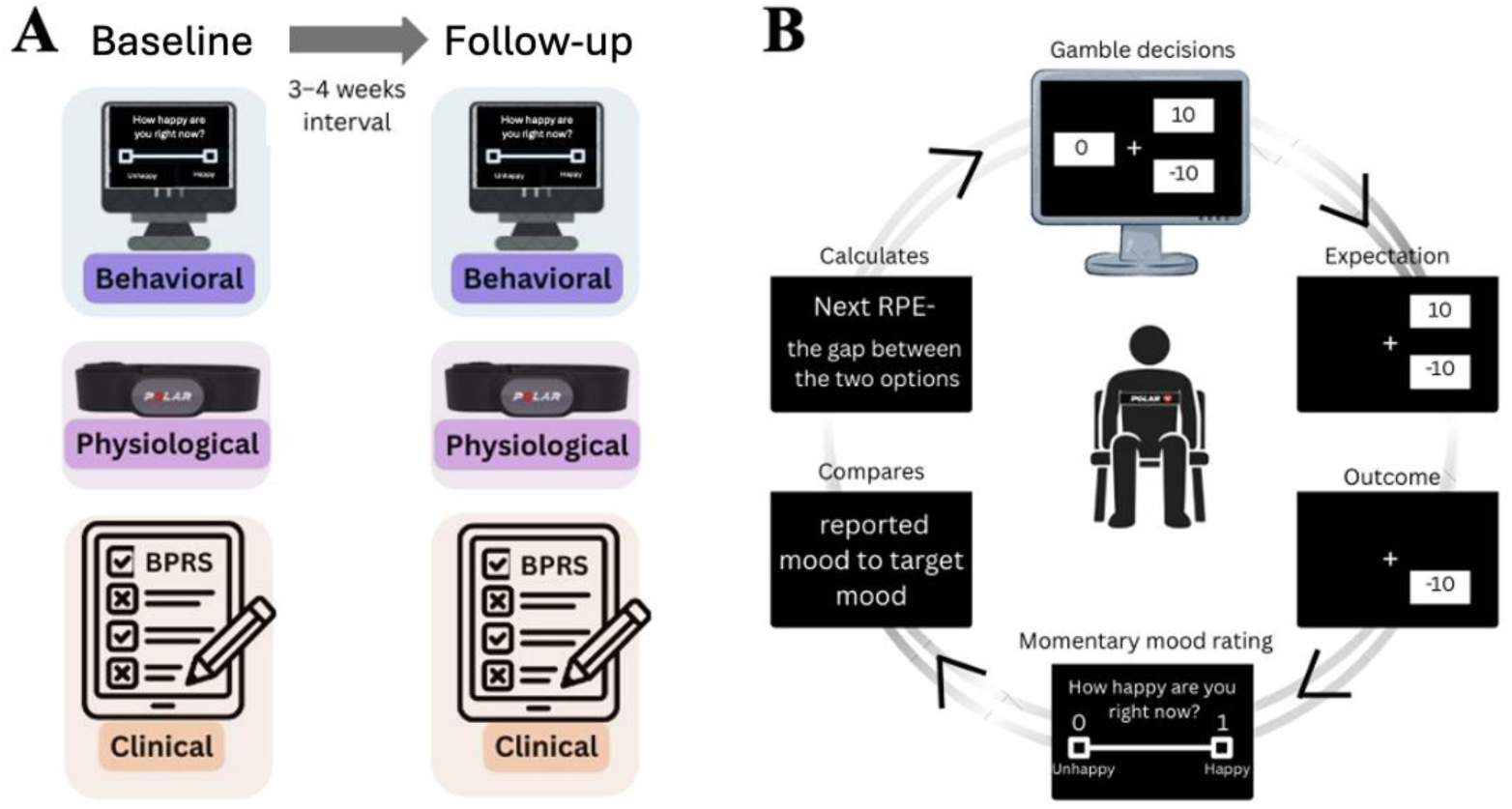
Study design. **(A)** The experimental protocol where patients completed two sessions during hospitalization. Each session included behavioral, physiological, and clinical assessments. **(B)** The experimental mood modification task: an adaptive reward-based task, where participants made repeated choices between a certain and a probabilistic gamble option (“gamble decisions”), forming expectations and receiving outcomes that generated reward prediction errors (RPEs). Participants periodically reported their subjective momentary mood, which was compared to a target mood state. The mood discrepancy from target state was used to adapt the subsequent RPEs, with the aim of modifying mood in the desired direction, upwards or downwards, thereby implementing a closed-loop modulation of emotional trajectories.

### Clinical assessment

Psychiatric symptom severity was assessed using the Brief Psychiatric Rating Scale (BPRS) (Overall & Gorham, 1962). The BPRS is a clinician-administered instrument that evaluates a broad range of psychiatric symptoms spanning affective, positive, negative, and general psychopathology domains. Total BPRS scores were used as an index of overall symptom severity.

In addition, symptom domain scores were examined to characterize associations between physiological, emotional, and clinical measures, following the five-factor structure proposed by Shafer (2005). Five symptom domains were derived: **Activation**, including Excitement (item 17), Tension (item 6), and Mannerisms and Posturing (item 7); **Resistance**, including Hostility (item 10), Suspiciousness (item 11), and Uncooperativeness (item 14); **Affect**, including Anxiety (item 2), Guilt Feelings (item 5), Depressive Mood (item 9), and Somatic Concern (item 1); **Positive Symptoms**, including Conceptual Disorganization (item 4), Hallucinatory Behaviour (item 12), Grandiosity (item 8), and Unusual Thought Content (item 15); and **Negative Symptoms**, including Emotional Withdrawal (item 3), Motor Retardation (item 13), and Blunted Affect (item 16). Domain scores were calculated as the mean of the constituent item scores. Clinical improvement was quantified as the change in total BPRS score between baseline and follow-up (ΔBPRS = BPRS₂ − BPRS₁), with more negative values indicating greater symptom improvement.

### Adaptive closed-loop mood modulation task

Participants completed an adaptive closed-loop reward-based decision making task designed to experimentally modulate momentary mood (Keren et al., 2021; Liuzzi et al., 2022; Jangraw et al., 2023). On each trial, participants chose between a certain outcome and a probabilistic gamble while periodically rating their current mood on a visual analogue scale ranging from *unhappy* to *happy* (Fig. 1B).

The task manipulated reward prediction errors (RPEs), defined as the discrepancy between expected and obtained outcomes. In this paradigm, RPEs were implemented by varying the difference between the two possible gamble outcomes, such that larger differences generated larger reward prediction errors. The task consisted of eight alternating mood blocks (four positive and four negative). Throughout the task, a closed-loop algorithm continuously monitored the discrepancy between the target mood for the current block and participants’ last reported mood. When participants remained far from the target mood, the algorithm progressively increased the magnitude of subsequent RPEs, thereby increasing the emotional impact of subsequent gains or losses. During positive blocks, larger positive RPEs were delivered to facilitate mood elevation, whereas during negative blocks more negative RPEs were delivered to facilitate mood reduction. To preserve uncertainty and maintain engagement, 30% of trials randomly delivered small outcomes opposite to the intended direction (loss in positive blocks and win in negative blocks). A detailed description of the task and adaptive algorithm have been reported previously (Keren et al., 2021; Liuzzi et al., 2022; Jangraw et al., 2023).

To quantify difficulty increasing positive mood, we used the cumulative positive RPE value during the positive mood blocks, when the adaptive algorithm provided increasingly larger positive RPEs to move mood toward the target. Higher values indicated that participants remained farther from the positive mood target and therefore required larger positive RPEs.

### Heart rate variability recording and preprocessing

Heart rate data was recorded continuously during the adaptive closed-loop mood modification task, using a Polar H10 chest strap. The device transmitted beat-to-beat interval (RR interval) data to the Elite HRV application, from which RR interval time series were exported. The Polar H10 has been validated against ECG for the assessment of RR intervals and HRV measures (Himariotis et al., 2022; Moya-Ramon et al., 2022; Vondrasek et al., 2023).

All HRV measures computed offline from the exported RR interval time series using custom Python scripts. First, RR interval series were preprocessed to reduce the influence of artefacts and ectopic beats. Physiologically implausible RR intervals (<300 ms or >2000 ms) were removed, and remaining artefacts were corrected using a local median-based procedure with a five-beat moving window. RR intervals deviating by more than 25% from the local median were replaced by the local median value, and the corrected RR interval series was subsequently linearly interpolated where necessary to preserve temporal continuity before spectral and nonlinear analyses, following current recommendations for HRV preprocessing (Vest et al., 2018).

Linear and nonlinear HRV measures were computed using different analysis durations to match their distinct methodological requirements. Linear time- and frequency-domain HRV measures were calculated from standardized 5-minute recordings, consistent with established recommendations for HRV assessment (Electrophysiology, 1996). The first chronologically recorded continuous RR segment lasting at least 8 minutes was selected; then the first 3 minutes were treated as a stabilization period, and the subsequent 5-minute segment was used for analysis (for one participant a pre-defined exception was applied, allowing a shorter 2.4-minute stabilization period). Nonlinear measures characterizing temporal complexity and dynamical behaviour generally benefit from longer time series and were therefore calculated from the longest available continuous RR recording for each participant to improve the stability of the measure (Rosenstein et al., 1993; Sassi et al., 2015). In the final HRV sample (57 patients and 29 healthy controls), nonlinear analysis duration ranged from 10.8 to 48.7 minutes (mean ± SD: 27.5 ± 6.9 minutes).

### Linear HRV measures

Linear time- and frequency-domain HRV measures were calculated from the artifact-corrected 5-minute RR interval series. The primary time-domain measure was the root mean square of successive differences (RMSSD), reflecting short-term parasympathetic modulation. Frequency-domain analysis was performed using Welch’s power spectral density estimation after interpolation of the RR interval series to a uniformly sampled signal (4 Hz). Low-frequency (LF; 0.04–0.15 Hz) and high-frequency (HF; 0.15–0.40 Hz) spectral power were calculated, and the LF/HF ratio was derived as an index of the relative distribution of autonomic oscillatory activity.

### Nonlinear HRV measures

To characterize the dynamic properties of cardiac regulation, nonlinear HRV measures were calculated from standardized RR interval series. The Lyapunov Exponent (LyapExp), which quantifies the exponential divergence of neighboring trajectories in reconstructed state space, providing an index of sensitivity to small perturbations in cardiac dynamics, was estimated using the Rosenstein (1993) method as implemented in nolds.lyap_r. The embedding dimension was fixed at *m* = 10, the trajectory length at 20 samples and τ = 1, whereas the lag and minimum temporal separation (min_tsep) were estimated automatically for each participant from the individual RR interval series. Approximate Entropy (ApEn), a measure of regularity and predictability of heartbeat fluctuations (Pincus, 1991), was calculated using parameters adapted to recording length according to predefined criteria implemented in the analysis pipeline. Recording-length-dependent implementation details are provided in the Supplementary Methods, and the parameters used for linear and nonlinear HRV analyses are summarized in Supplementary Table S1.

### Medication load

To account for potential medication-related effects on autonomic measures, overall medication burden was quantified using a normalized dose index. For each prescribed medication, the reported daily dose (mg/day) was divided by a reference minimum effective dose obtained from drug labelling information and relevant clinical literature. The resulting normalized values were summed across all medications prescribed to each participant, yielding a composite medication load score. This index included both psychiatric and non-psychiatric medications and was entered as a covariate in all regression models examining subsequent clinical outcome.

### Statistical analyses

Statistical analyses were conducted in Python using standard scientific libraries, including NumPy, SciPy, pandas, and statsmodels.

Group differences between patients and healthy controls in behavioural and physiological measures were assessed using Welch’s *t*-tests because of unequal sample sizes and heterogeneity of variances. Effect sizes are reported as Cohen’s *d*. Associations between behavioural, autonomic, and clinical variables were examined using Pearson correlation coefficients.

Hierarchical multiple linear regression analyses were performed to evaluate prospective associations between baseline emotional and autonomic measures and subsequent symptom improvement. **Clinical improvement** was first quantified as the change in symptom severity between baseline and follow-up (ΔBPRS = BPRS₂ − BPRS₁), with more negative values indicating greater symptom improvement. As a complementary analysis, clinical improvement was also quantified using a residualized improvement score. Specifically, ΔBPRS was regressed on baseline symptom severity (BPRS₁), and the residuals from this regression were used as the outcome measure, representing symptom improvement after accounting for individual differences in baseline clinical severity.

A baseline model including age, sex, medication load, and baseline symptom severity (BPRS₁) was first constructed. Individual emotional and autonomic measures were subsequently evaluated by adding each measure separately to the baseline model. These measures included the emotional measure of difficulty increasing mood, derived from positive reward prediction error (RPE Positive) trials, and the autonomic linear HRV measures RMSSD and LF/HF ratio, as well as the nonlinear HRV measures Approximate Entropy (ApEn) and Lyapunov Exponent (LyapExp).

Because LF/HF ratio, LyapExp, and ApEn each showed individual prospective associations with subsequent symptom change, these three measures were then combined to test whether their complementary information could be well-represented by a single multidimensional autonomic index. Two composite HRV indices were evaluated. The primary composite autonomic index was calculated by first z-standardizing three directionally aligned HRV measures within the fixed common prediction sample: inverted LF/HF ratio (−LF/HF), LyapExp and ApEn. **The composite score** was then defined as the unweighted arithmetic mean of these three standardized measures, such that higher scores reflected lower LF/HF, higher LyapExp and higher ApEn. A second alternative composite index was derived using principal component analysis (PCA) applied to the same three standardized measures.

The robustness of the regression findings was evaluated using bootstrap resampling (10,000 iterations) and leave-one-participant-out sensitivity analyses. Test–retest reliability of the autonomic measures and composite score across sessions was assessed using intraclass correlation coefficients (ICC; two-way mixed-effects model, single measurement; ICC [3,1]). Statistical significance was defined as *p* < 0.05.

## Results

### Psychiatric patients show altered affective reactivity and autonomic dynamics

Emotional and autonomic measures differed between psychiatric patients and healthy controls (Fig. 2). Patients showed lower mood reactivity to positive rewards, as reflected by greater difficulty increasing mood during positive blocks (714.38 ± 304.64 vs. 335.41 ± 185.51, *p* < 0.001, Cohen’s *d* = 1.33). They also showed lower RMSSD (20.97 ± 15.01 vs. 28.76 ± 12.78 ms, *p* = 0.014, *d* = −0.54), and higher LF/HF ratio (5.66 ± 5.59 vs. 2.42 ± 2.37, *p* < 0.001, *d* = 0.68). Patients also showed alterations in nonlinear HRV measures, specifically lower LyapExp (0.0284 ± 0.0169 vs. 0.0375 ± 0.0122, *p* < 0.01, *d* = −0.58) and lower ApEn (1.10 ± 0.36 vs. 1.39 ± 0.27, *p* < 0.001, *d* = −0.86).

**Figure 2.**
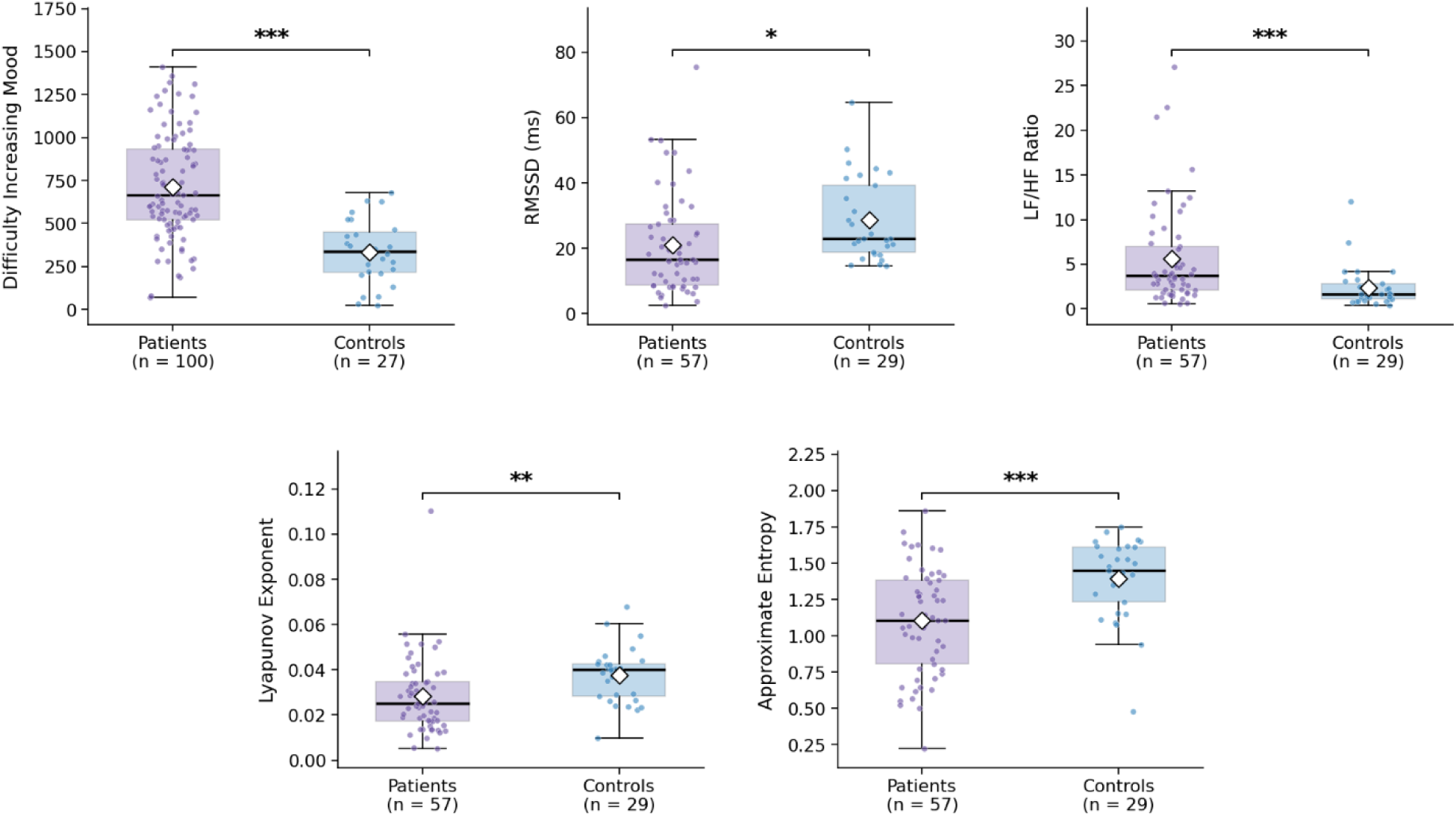
Emotional and autonomic measures in psychiatric patients versus healthy controls. Patients showed greater difficulty increasing mood during positive blocks, lower RMSSD, LyapExp and ApEn, and higher LF/HF ratio compared to healthy controls. Boxplots show medians and interquartile ranges; whiskers indicate 1.5×IQR, points represent individual participants, and diamonds indicate group means. Group differences were assessed using two-sided Welch’s t-tests. Sample sizes differ across panels because HRV analyses were restricted to participants with high-quality physiological recordings (* indicates p<0.05, ** p<0.01, and *** p<0.001).

### Associations with current symptom dimensions

Among the measures that differentiated patients from healthy controls, three showed initial nominal associations with clinical symptom dimensions in the patient sample (Fig. 3). Greater difficulty increasing mood was associated with greater affective symptom severity (*r* = 0.33, *p* < 0.001), as well as higher resistance (*r* = 0.24, *p* = 0.015) and activation symptoms (*r* = 0.25, *p* = 0.011; Fig. 3a and Fig. 4a–c). Neither RMSSD nor LF/HF ratio was significantly associated with any clinical symptom dimension (Fig. 3b,c), but lower ApEn was associated with greater negative symptom severity (*r* = −0.36, *p* = 0.006) and higher overall BPRS scores (*r* = −0.27, *p* = 0.043; Fig. 3e and Fig. 4d,e), whereas higher LyapExp was associated with greater positive symptom severity (*r* = 0.39, *p* = 0.003), negative symptom severity (*r* = 0.29, *p* = 0.028), and resistance (*r* = 0.29, *p* = 0.027, Fig. 3d).

**Figure 3.**
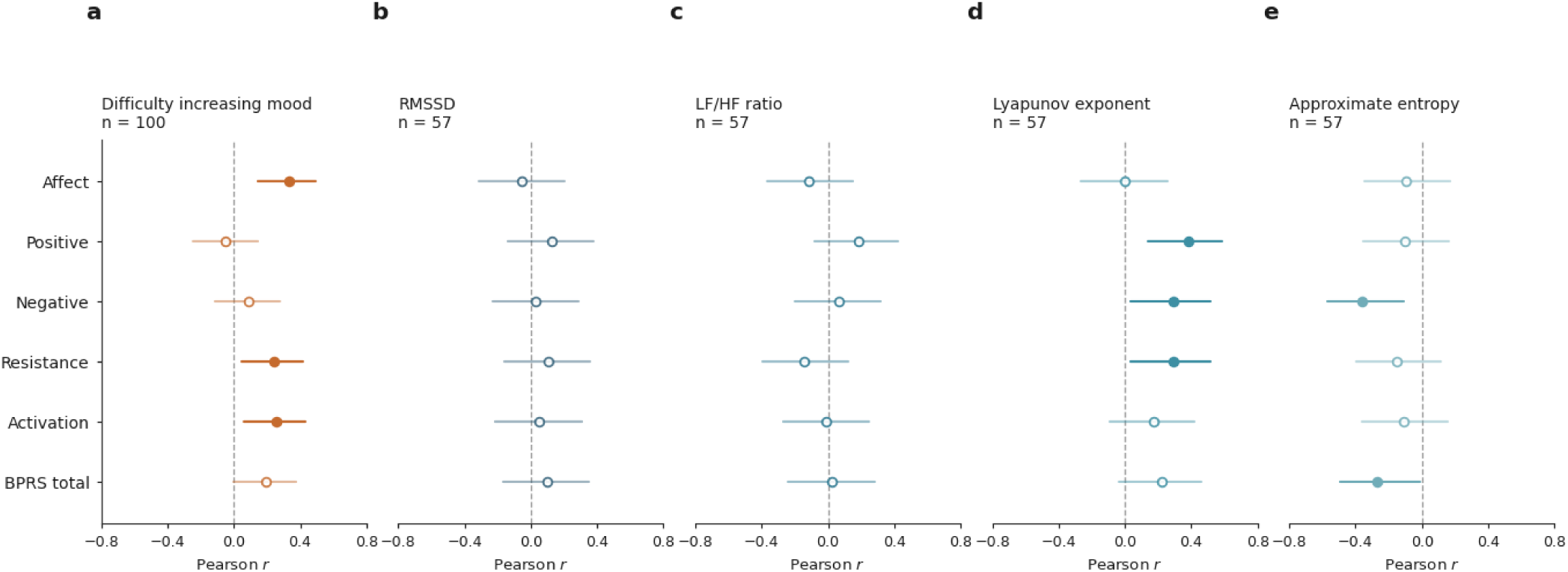
Nominal associations of emotional reactivity and autonomic measures across clinical symptom dimensions. Pearson correlations between difficulty increasing mood (a), RMSSD (b), LF/HF ratio (c), LyapExp (d) and ApEn (e) and clinical symptom dimensions in psychiatric patients. Each circle indicates correlations to a different clinical domain. Distance of circles from the dashed vertical line indicates Pearson correlation coefficients and the line itself denotes *r* = 0. Filled circles indicate nominally significant associations (*p* < 0.05), whereas open circles indicate non-significant associations. Difficulty increasing mood analyses included *n* = 100 participants, whereas autonomic analyses included *n* = 57 participants.

**Figure 4.**
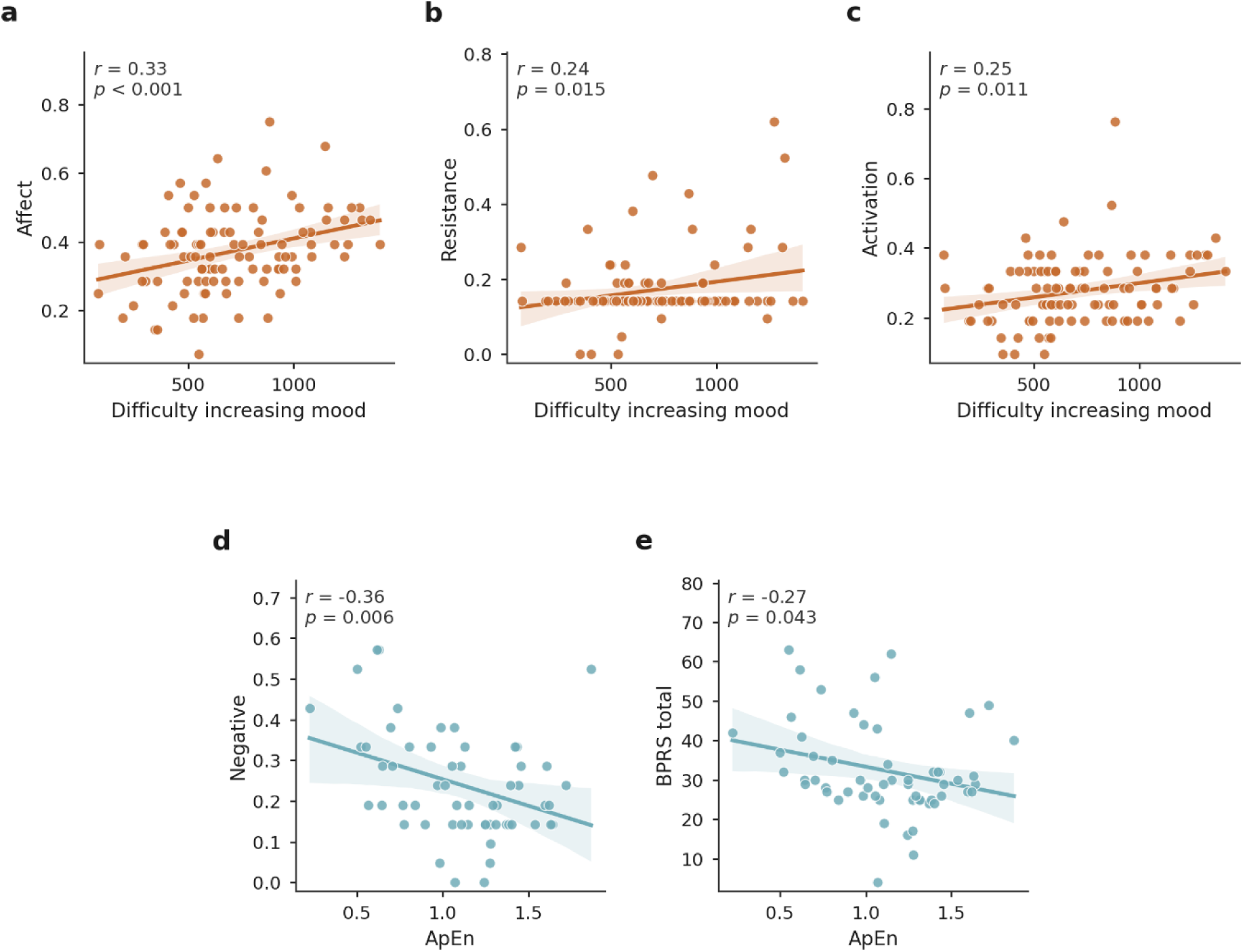
Associations with clinical symptom dimensions after outlier sensitivity analysis. Scatterplots illustrating selected associations between difficulty increasing mood and affective (a), resistance (b) and activation (c) symptom severity, and between ApEn and negative symptom severity (d) and total BPRS score (e). Points represent individual participants. Solid lines indicate linear regression fits and shaded areas indicate 95% confidence intervals. Pearson correlation coefficients (*r*) and two-sided *p* values are shown within each panel. Difficulty increasing mood analyses included *n* = 100 participants and ApEn analyses included *n* = 57 participants.

However, the clinical associations involving LyapExp were all sensitive to exclusion of a single highest outlier observation (*z* = 4.86). Following exclusion of this participant, the associations were attenuated and no longer significant for positive symptoms (*r* = 0.11, *p* = 0.411), negative symptoms (*r* = 0.10, *p* = 0.481), or resistance (*r* = 0.19, *p* = 0.172; Supplementary Fig. S1 and Supplementary Table S3).

After correction for multiple comparisons, only the associations between difficulty increasing mood and affective symptoms (FDR-adjusted *q* = 0.023) and between LyapExp and positive symptoms (FDR-adjusted *q* = 0.046) remained significant in the full sample.

Thus, the association between difficulty increasing mood and affective symptom severity was the only cross-sectional symptom association that was robust to both multiple-comparisons correction and outlier sensitivity analysis.

### Prospective associations with subsequent clinical improvement

Among the 59 participants with clinical BPRS assessments available at both baseline and follow-up, mean symptom severity decreased from 33.19 ± 8.67 to 29.25 ± 8.89, corresponding to a mean ΔBPRS of −3.93 ± 9.78. Despite this overall mean improvement, clinical trajectories varied considerably (Fig 5): 39 participants (66.1%) showed symptom improvement (ΔBPRS < 0), whereas 20 (33.9%) showed no improvement (ΔBPRS ≥ 0). The prospective analysis sample comprised 34 of these participants, who had both valid baseline HRV recordings and complete clinical assessments at follow-up. In this subsample, clinical improvement was similar to that observed in the larger longitudinal sample: mean BPRS scores decreased on average from 34.62 ± 9.99 at baseline to 30.00 ± 9.89 at follow-up, with mean ΔBPRS = −4.62 ± 11.59; 22 participants (64.7%) improved, whereas 12 (35.3%) showed no improvement. This variability in clinical trajectories underscores the need for baseline measures that can be informative about individual differences in treatment response.

**Figure 5.**
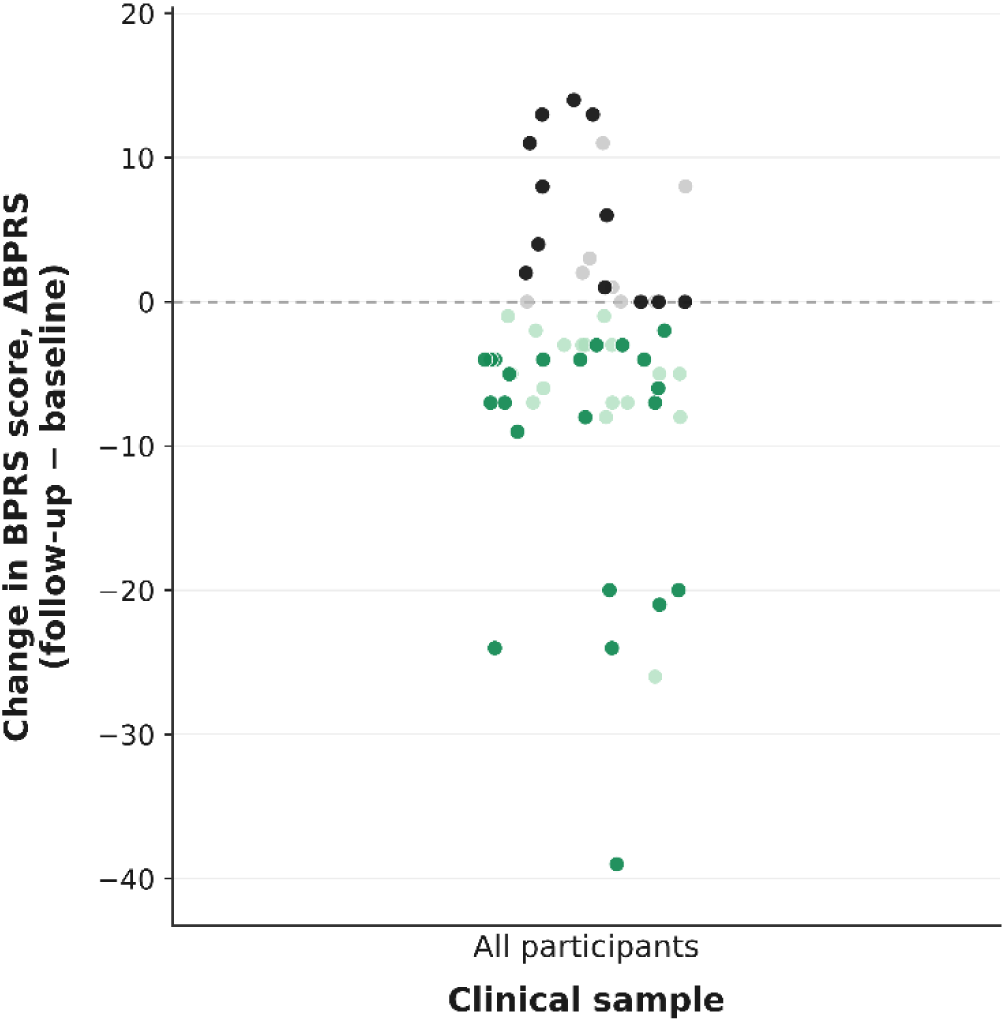
Distribution of symptom change in the longitudinal clinical sample. Each point represents one participant with BPRS assessments available at baseline and follow-up (n = 59). ΔBPRS was calculated as the follow-up minus baseline score; negative values indicate symptom improvement, whereas values of zero or greater indicate no improvement. Participants included in the prospective analysis sample (n = 34) are shown in dark green (improvement) or black (no improvement), whereas participants outside the prospective analysis are shown in light green or grey, respectively. Points are horizontally jittered for visual clarity.

To determine whether baseline emotional and autonomic measures can provide additional prognostic information, each candidate measure was added separately to the clinical baseline model, which included the baseline BPRS score, age, sex and medication load. This approach enabled evaluation of the unique contribution of each measure beyond the baseline model, which was maintained as a reference model across analyses (standardized coefficients, 95% confidence intervals, *t* statistics and *p* values for the clinical and tested measures are reported in Supplementary Table S4).

The emotional reactivity measure did not significantly improve the model beyond the clinical baseline model (β = −0.082, *p* = 0.535), accounting for only an additional 0.6% of the explained variance (*R²* = 0.562, adjusted *R²* = 0.484; Δ*R²* = 0.006; ΔAIC = +1.52). Similarly, RMSSD was not significantly associated with subsequent symptom change after adjustment for the clinical baseline variables (β = −0.218, *p* = 0.096; *R²* = 0.599, adjusted *R²* = 0.527; Δ*R²* = 0.043; ΔAIC = −1.43).

Among the other autonomic measures, the LF/HF ratio showed a significant incremental associaition beyond the clinical baseline model (β = 0.304, *p* = 0.028; *R*² = 0.628, adjusted *R*² = 0.561; Δ*R*² = 0.072; ΔAIC = −3.99). LyapExp was also significantly associated with subsequent symptom change beyond the baseline model (β = −0.297, *p* = 0.037; *R*² = 0.621, adjusted *R*² = 0.553; Δ*R*² = 0.065; ΔAIC = −3.36). ApEn similarly showed a significant association (β = −0.269, *p* = 0.046; *R*² = 0.616, adjusted *R*² = 0.547; Δ*R*² = 0.060; ΔAIC = −2.90). Together, these findings indicate that higher LF/HF ratio, lower LyapExp, and lower ApEn at baseline were each associated with less subsequent clinical improvement after accounting for baseline clinical state and characteristics.

Because LF/HF ratio, LyapExp, and ApEn each showed individual prospective associations with subsequent symptom change, we next asked whether integrating these complementary autonomic features into a single index could capture their convergent relationship with clinical improvement more consistently than any individual measure alone.

LF/HF ratio, LyapExp, and ApEn were combined into a Mean-Z autonomic composite. The composite showed the largest incremental association beyond the baseline model with symptom improvement (*F* change(1,28) = 10.30, *p* = 0.003), increasing the explained variance by 11.9% (*R*² = 0.675, adjusted *R*² = 0.618; Δ*R*² = 0.119; ΔAIC = −8.65). The composite index showed a stronger association with subsequent symptom change than any individual HRV measure alone (β = -0.390, *p* = 0.003), supporting the potential value of integrating complementary autonomic information (Fig. 6; Supplementary Table S5).

**Figure 6.**
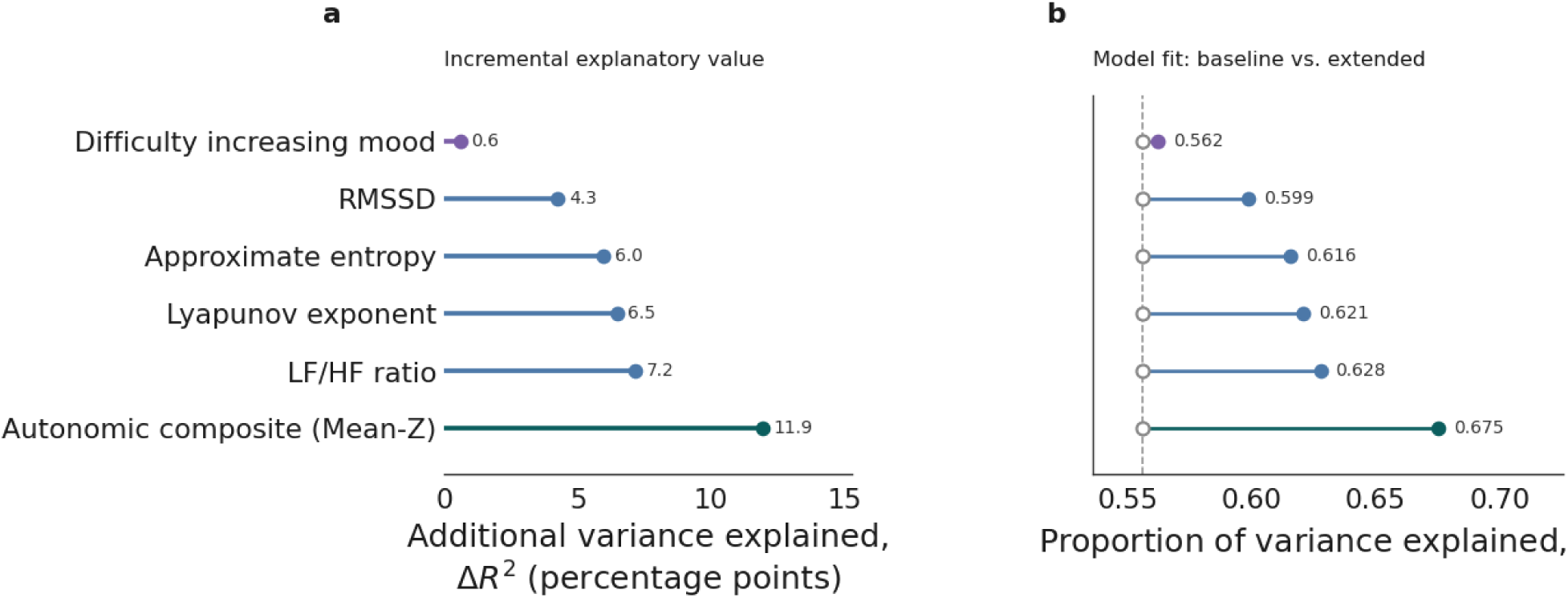
Association of emotional and autonomic measures with clinical improvement. **(a)** The additional variance explained (Δ*R*²) by each emotional or autonomic predictor when added separately to the clinical baseline model. (**b)** Proportion of variance explained (*R*²) by the clinical baseline model and each corresponding extended model. The clinical baseline model included baseline BPRS score, age, sex and medication load. The dashed vertical line indicates the variance explained by the clinical baseline model (*R*² = 0.556).

To determine whether the observed association reflected more than the well-established relationship between baseline symptom severity and subsequent improvement, we repeated the analyses using a residualized clinical improvement score as the outcome measure. This outcome quantifies symptom change relative to the expected based on baseline symptom severity. The composite autonomic index remained significantly associated with treatment response under this alternative outcome definition (β = -0.420, *p* = 0.011; R² = 0.435), indicating that the association was not solely driven by baseline symptom severity.

We next examined whether the findings depended on the method used to construct the autonomic composite. A PCA-derived composite yielded virtually identical results (β = -0.390, *p* = 0.004; R² = 0.674), accounting for a comparable increase in explained variance beyond the clinical baseline model (ΔR² = 0.118). These findings indicate that the prospective association was robust to the method used to derive the composite autonomic index.

Test-retest reliability of the autonomic composites index was evaluated in 31 participants who had HRV data available at both assessment sessions. The two composite indices showed comparable, modest test–retest reliability: ICC(3,1) = 0.481 (*p* = 0.003) for the Mean-Z composite and ICC(3,1) = 0.489 (*p* = 0.002) for the PCA-derived composite.

To further evaluate the robustness of the findings, we performed a bootstrap resampling analysis with 10,000 iterations. For the primary treatment-response outcome (ΔBPRS), the Mean-Z composite yielded a median standardized coefficient of β = -0.378 (95% CI [-0.695, - 0.119]), with the direction of the association preserved in 99.7% of bootstrap samples. Using the residualized clinical improvement outcome produced similar results (median β = −0.399, 95% CI [−0.815, −0.066]), with the expected direction of the association preserved in 99.2% of bootstrap samples.

Finally, we assessed the influence of individual participants using a leave-one-participant-out sensitivity analysis. For the primary ΔBPRS outcome, standardized coefficients ranged from β = -0.435 to β = -0.272, with statistical significance retained in all iterations (100%). Similar results were obtained for the residualized clinical improvement outcome (β range = −0.527 to −0.279), with all coefficients retaining the expected direction and statistical significance retained in 33 of 34 iterations (97.1%). Collectively, these analyses indicate that the prospective association of the autonomic composite index was robust across alternative outcome definitions, alternative composite construction methods, bootstrap resampling, and systematic exclusion of individual participants (although the residualized model became non-significant following the exclusion of one participant).

**Table 2.** Summary of robustness and validation analyses for the autonomic composite index.

| Validation analysis | Main result | Purpose |
| --- | --- | --- |
| Mean-Z Composite Model | $\beta = -0.390$, $p = 0.003$, $R^2 = 0.675$ | Main model |
| PCA Composite Model | $\beta = -0.390$, $p = 0.004$, $R^2 = 0.674$ | Alternative composite construction |
| Residualized clinical improvement | $\beta = -0.420$, $p = 0.011$, $R^2 = 0.435$ | Alternative outcome definition |
| Test-retest reliability | ICC (3,1) = 0.481 ($p = 0.003$; Mean-Z);<br>ICC (3,1) = 0.489 ($p = 0.002$; PCA) | Temporal stability |
| Bootstrap (10,000 resamples) | Median $\beta = -0.378$, 95% CI [-0.695, -0.119], 99.7% in the expected direction | Resampling validation |
| Leave-one-participant-out | $\beta$ range = $-0.435$–$-0.272$, significant in 100% of iterations | Sensitivity analysis |

**Note.** The Mean-Z composite was calculated as the unweighted mean of z-standardized $-LF/HF$ , $LyapExp$ and $ApEn$ values, such that higher scores reflected lower $LF/HF$ , higher $LyapExp$ and higher $ApEn$ . The PCA composite represents the first principal component derived from the same directionally aligned standardized measures. Residualized clinical improvement was defined as standardized ΔBPRS residualized for baseline BPRS severity. Test-retest reliability was evaluated in the 31 participants with HRV measurements available at both assessment sessions. Bootstrap analyses were based on 10,000 resamples. $\beta$ values represent standardized regression coefficients.

## Discussion

The present study examined whether baseline emotional reactivity and autonomic measures could distinguish psychiatric patients from healthy individuals, capture clinically meaningful variation in symptom severity, and provide prospective information about subsequent clinical change during hospitalization. By combining a task-derived measure of positive emotional reactivity, with time-domain, frequency-domain, and nonlinear measures of heart rate variability (HRV), we assessed dimensions of emotional and physiological regulation in a transdiagnostic clinical sample. The findings suggest that these measures have partly distinct clinical relevance: mood reactivity was most strongly related to concurrent affective symptoms, whereas autonomic HRV measures - particularly when integrated into a composite index - provided the most consistent prospective association with clinical improvement. Together, these findings suggest that emotional reactivity and autonomic dynamics may provide temporally complementary information about current symptom expression versus subsequent clinical change, potentially reflecting distinct dimensions of psychiatric functioning.

Because the present task adaptively manipulated reward outcomes to influence mood, difficulty increasing mood does not simply reflect lower average mood; rather, it captures the extent to which mood does not increase in response to favorable outcomes even as those outcomes become progressively more positive, providing a task-based measure of positive affective reactivity. This finding is consistent with evidence that psychiatric symptoms, particularly depression and anhedonia, are characterized by diminished responsiveness to reward and reduced effects of positive outcomes on momentary affect (Cléry-Melin et al., 2019; Horne et al., 2021; Rutledge et al., 2017). Computational and neuroimaging studies have further linked depressive and psychotic symptoms to altered reward-prediction-error signalling and attenuated recruitment of corticostriatal reward systems (Gradin et al., 2011; Keren et al., 2018; Sharma et al., 2017).

The association between difficulty increasing mood and affective symptom severity, which remained significant after correction for multiple comparisons, further supports the clinical relevance of this measure. In contrast, positive affective reactivity was not prospectively associated with subsequent clinical change beyond baseline symptom severity, age, sex, and medication load. This pattern suggests that the measure may be more closely related to patients’ current affective state than to their subsequent clinical change – highlighting that measures informative about current symptom expression are not necessarily those that provide prospective information about longitudinal clinical trajectory.

Patients also differed from healthy controls across the autonomic measures examined. Relative to controls, patients showed lower RMSSD, higher LF/HF ratios, lower ApEn, and lower LyapExp. These findings are broadly consistent with extensive evidence of altered cardiac autonomic regulation across depressive, anxiety-related, and psychotic disorders (Alvares et al., 2016; Clamor et al., 2016; Koch et al., 2019; Z. Wang et al., 2025). Importantly, the largest autonomic effect sizes were observed for the nonlinear measures. ApEn quantifies the regularity and predictability of fluctuations within a time series (Pincus, 1991), whereas LyapExp, estimated using a Rosenstein-based approach, characterizes the rate at which initially neighbouring trajectories diverge within a reconstructed dynamical system (Rosenstein et al., 1993). These measures therefore capture distinct, though potentially related, properties of autonomic organization, and their concurrent alteration should not be interpreted as interchangeable indicators of a single construct such as “reduced complexity.” Rather, the findings suggest broader alteration in the temporal organization of cardiac regulation, encompassing both sequential regularity and sensitivity to local dynamical perturbations. This is consistent with previous work showing that nonlinear cardiac dynamics can reveal autonomic alterations not captured by conventional linear measures in both schizophrenia and in depression (Bär et al., 2007; Schulz et al., 2010; Valenza et al., 2015).

The cross-sectional symptom findings further illustrated the distinct clinical roles of the emotional and autonomic measures. Difficulty increasing mood was primarily associated with affective symptoms, whereas the nonlinear autonomic indices showed nominal associations with negative symptoms and overall psychopathology — broadly consistent with reports linking reduced cardiac complexity to depression, schizophrenia, and shifts in affective state (Bär et al., 2007; Gentili et al., 2017; Valenza et al., 2015; Wazen et al., 2018). In contrast, RMSSD and LF/HF distinguished patients from healthy controls but were not significantly associated with symptom dimensions within the patient group. Thus, the ability of a measure to detect a group-level difference does not necessarily imply that it tracks variation in symptom severity within a clinical sample. The ApEn associations did not survive correction for multiple comparisons and should therefore be regarded as tentative, whereas the Lyapunov-related associations were not robust to a sensitivity analysis excluding one extreme observation and were no longer statistically significant following its removal. The Lyapunov findings should therefore not be treated as evidence of a symptom-specific relationship. Cross-sectional associations between autonomic measures and symptom severity were comparatively weak and inconsistent, suggesting that their clinical relevance may not primarily lie in tracking concurrent symptom burden.

In contrast, the prospective analyses revealed that LF/HF ratio, LyapExp, and ApEn were each associated with subsequent clinical improvement. Integrating these complementary autonomic measures yielded a stronger and more consistent association with clinical change than any individual physiological measure alone. Previous studies have similarly suggested that integrating complementary HRV features may provide more informative clinical characterization than relying on a single measure (Byun et al., 2019; Chang et al., 2009; Stout et al., 2022), consistent with broader work showing that integration across physiological features can improve characterization of clinical trajectories in psychiatric populations (Habets et al., 2023). Because LF/HF, ApEn, and LyapExp quantify different properties of cardiac regulation, their integration may provide a broader representation of autonomic organization while reducing dependence on the measurement noise and conceptual limitations of any single index. Importantly, the association was reproduced when the composite was constructed using principal component analysis, when clinical change was represented by a residualized outcome accounting for baseline symptom severity, and across bootstrap and leave-one-participant-out analyses. The association was preserved when the composite was constructed using principal component analysis, when clinical change was represented by a residualized outcome accounting for baseline symptom severity, and across bootstrap and leave-one-participant-out analyses. This consistency reduces the likelihood that it depended exclusively on a particular method of composite construction, definition of clinical change, nevertheless, these results should be viewed as preliminary prognostic evidence only given the modest sample size and the absence of external validation.

At a theoretical level, the findings are broadly compatible with the neurovisceral integration model (Thayer & Lane, 2000), which proposes that cardiac vagal regulation reflects central– peripheral mechanisms supporting attentional and emotional regulation, such that reduced vagally mediated HRV is associated with poorer self-regulation and reduced behavioural flexibility. Reduced affective reactivity in response to positive outcomes and prediction errors and altered autonomic organization may therefore both reflect limitations in adaptating to changing internal and environmental conditions. The emotional task may be more sensitive to context-dependent affective responding, whereas HRV-derived measures may capture broader features of autonomic organization. The results therefore support a multidimensional view of emotional-autonomic regulation in which related but distinguishable processes may carry temporally distinct information about current affective symptoms and subsequent clinical change.

Several limitations should be noted. The prognostic analyses were based on a modest sample (*n* = 34), limiting the precision and generalizability of the estimates despite stable directionality across bootstrap and leave-one-participant-out analyses. The composite indices showed only modest test–retest reliability, limiting confidence that they reflect reliable trait-like individual differences. The linear measures were derived from standardized 5-minute windows, whereas the nonlinear indices were estimated from the longest available continuous recording (10.8– 48.7 minutes), which may have affected measurement precision and comparability across indices. The transdiagnostic sample, while consistent with a dimensional approach to psychiatric heterogeneity (Cuthbert & Insel, 2013; Leucht et al., 2024), was heterogeneous in diagnosis, medication exposure, and clinical course, limiting disorder-specific inference. The observational design precludes causal conclusions. Finally, the autonomic composite index was constructed after identifying the individual autonomic measures associated with clinical change and was evaluated in the same longitudinal sample. Its stronger association should therefore be interpreted as an integrative summary of convergent within-sample findings rather than as independent validation of a prognostic biomarker. The clinical relevance of this multidimensional profile will need to be validated in larger independent samples using a prespecified processing and modelling pipeline.

Despite these limitations, the measures examined here may have practical value because they can be derived from non-invasive, accessible, and relatively inexpensive cardiac recordings, including wearable devices (L. Wang et al., 2024). If replicated, autonomic measures could complement clinical assessment by providing information not fully captured by baseline symptom severity. At the same time, the measurement accuracy of consumer-grade wearables is not always equivalent to that of clinical-grade ECG recording, and further validation of HRV indices - particularly the nonlinear measures, which may be more sensitive to signal noise - is needed before such applications can be considered.

In conclusion, baseline affective reactivity and autonomic measures provided complementary information about psychiatric status and subsequent clinical change. Positive affective reactivity was more robustly related to concurrent affective symptoms, whereas several autonomic measures were prospectively associated with subsequent symptom improvement. Integrating these individually informative autonomic features into a composite index yielded a stronger and more consistent relationship with clinical change than any single measure alone. Although independent replication is required, these findings suggest that multidimensional autonomic dynamics may provide accessible physiological information about psychiatric clinical trajectories that is not captured by baseline symptom severity alone.

## Data Availability

De-identified data supporting the findings of this study are available from the corresponding author upon reasonable request, subject to ethical and institutional approval.

## Appendices

**Supplementary Table S1.** |Implementation parameters for linear and nonlinear HRV analyses.

| Measure | Method | Parameters |
| --- | --- | --- |
| RMSSD | Standard | - |
| LF/HF | Welch PSD | fs=4 Hz; LF 0.04–0.15; HF 0.15–0.40 |
| ApEn | Pincus | $m = 2-3$; $r = 0.15-0.20 \times \text{SD}$, selected according to recording length |
| LyapExp | Rosenstein<br>(nolds.lyap_r) | $m = 10$; $\tau = 1$; trajectory length = 20; lag and min_tsep estimated automatically |

**Note:** LF/HF was estimated using Welch's power spectral density (PSD) method with a sampling frequency of 4 Hz; LF and HF bands were defined as 0.04–0.15 Hz and 0.15–0.40 Hz, respectively. For ApEn, $m = 3$ and $r = 0.15 \times \text{SD}$ were used for recordings containing >3,000 RR intervals, whereas $m = 2$ and $r = 0.20 \times \text{SD}$ were used for shorter recordings. LyapExp was estimated from standardized RR interval series using the Rosenstein method implemented in nolds.lyap\_r, with embedding dimension $m = 10$ , $\tau = 1$ and trajectory length = 20; lag and minimum temporal separation (min\_tsep) were estimated automatically for each participant.

**Supplementary Table S2.** Sensitivity analysis excluding the highest LyapExp value.

| Sample | Association | Full sample |  |  |  | Highest LyapExp value excluded |  |  |  |
| --- | --- | --- | --- | --- | --- | --- | --- | --- | --- |
|  |  | n | r | 95% CI | p | n | r | 95% CI | p |
| Patients | RMSSD–LyapExp | 57 | 0.42 | [0.18, 0.62] | 0.001 | 56 | 0.47 | [0.24, 0.65] | <0.001 |
| Patients | LF/HF–LyapExp | 57 | -0.41 | [-0.60, -0.17] | 0.002 | 56 | -0.44 | [-0.63, -0.20] | <0.001 |
| Patients | LyapExp–ApEn | 57 | 0.08 | [-0.19, 0.33] | 0.566 | 56 | 0.26 | [0.00, 0.49] | 0.050 |
| Combined sample | RMSSD–LyapExp | 86 | 0.41 | [0.22, 0.57] | <0.001 | 85 | 0.45 | [0.26, 0.60] | <0.001 |
| Combined sample | LF/HF–LyapExp | 86 | -0.44 | [-0.60, -0.25] | <0.001 | 85 | -0.47 | [-0.62, -0.29] | <0.001 |
| Combined sample | LyapExp–ApEn | 86 | 0.22 | [0.01, 0.41] | 0.043 | 85 | 0.38 | [0.18, 0.55] | <0.001 |

**Note:** Values are two-sided Pearson correlation coefficients. 95% confidence intervals were calculated using Fisher’s z transformation. The sensitivity analysis excluded the participant with the highest LyapExp value from the patient and combined-sample analyses only; the control-only analysis was not altered because the extreme observation occurred in the patient group. The participant was retained in the primary analyses. No multiple-comparison correction was applied to these exploratory inter-autonomic correlations.

**Supplementary Figure S1.**
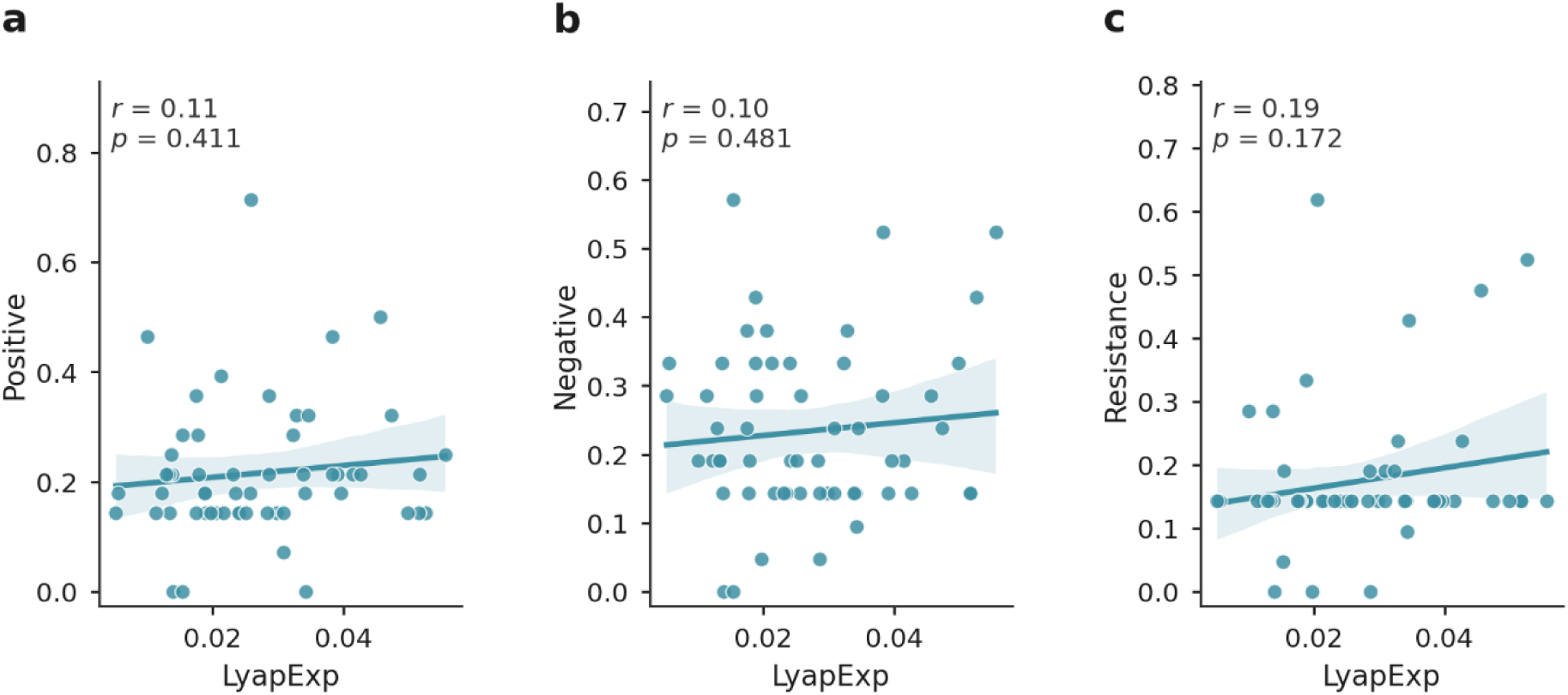
Sensitivity of LyapExp–symptom associations to exclusion of an outlier LyapExp value. Associations between LyapExp and positive (a), negative (b) and resistance (c) symptom dimensions after exclusion of the participant with the highest LyapExp value. Points represent individual participants, solid lines indicate linear regression fits and shaded areas indicate 95% confidence intervals. Pearson correlation coefficients (*r*) and two-sided *p* values are shown within each panel. Following exclusion of the extreme observation, all three associations were attenuated and were no longer statistically significant.

**Supplementary Table S3.** Sensitivity of LyapExp associations with clinical symptom dimensions to exclusion of the highest LyapExp value.

| Clinical symptom dimension | Full sample |  |  |  |  | Highest LyapExp value excluded |  |  |  |
| --- | --- | --- | --- | --- | --- | --- | --- | --- | --- |
|  | n | r | 95% CI | p | FDR-adjusted q | n | r | 95% CI | p |
| **Positive** | 57 | 0.39 | [0.14, 0.59] | 0.003 | 0.046 | 56 | 0.11 | [-0.16, 0.36] | 0.411 |
| **Negative** | 57 | 0.29 | [0.03, 0.51] | 0.028 | 0.119 | 56 | 0.10 | [-0.17, 0.35] | 0.481 |
| **Resistance** | 57 | 0.29 | [0.04, 0.51] | 0.027 | 0.119 | 56 | 0.19 | [-0.08, 0.43] | 0.172 |

**Note:** Values are two-sided Pearson correlation coefficients. 95% confidence intervals were calculated using Fisher's z transformation. FDR-adjusted q values refer to the full set of 30 primary emotional/autonomic-clinical correlations and were computed using the Benjamini-Hochberg procedure. The sensitivity analysis excluded the participant with the maximum LyapExp value (0.11049), which was 4.86 sample standard deviations above the LyapExp mean and also exceeded the Tukey upper fence. This exclusion was performed only as a sensitivity analysis; the participant was retained in the primary analyses. FDR correction was not recomputed after exclusion.

**Supplementary Table S4.** Standardized effects of the emotional and autonomic predictors of clinical improvement.

| Predictor | Standardized $\beta$ | CI 95% | t | p |
| --- | --- | --- | --- | --- |
| **Baseline BPRS** | -0.408 | [-0.728, -0.089] | -2.615 | 0.014* |
| **RPE pos** | -0.082 | [-0.350, 0.186] | -0.627 | 0.535 |
| **RMSSD** | -0.217 | [-0.477, 0.041] | -1.724 | 0.096 |
| **LF/HF** | 0.304 | [0.036, 0.573] | 2.321 | 0.028* |
| **LyapExp** | -0.297 | [-0.575, -0.019] | -2.187 | 0.037* |
| **ApEn** | -0.269 | [-0.534, -0.005] | -2.083 | 0.046* |
| **Mean-Z composite** | -0.390 | [-0.639, -0.141] | -3.210 | 0.003** |

**Supplementary Table S5.** Comparison of models relating baseline measures to subsequent change in BPRS scores.

| Model | R<sup>2</sup> | Adj. R<sup>2</sup> | AIC | Added predictor (β) | p |
| --- | --- | --- | --- | --- | --- |
| **Baseline model** | 0.556 | 0.495 | 244.50 | - | - |
| **+RPE** | 0.562 | 0.484 | 246.02 | -0.082 | 0.535 |
| **+RMSSD** | 0.599 | 0.527 | 243.07 | -0.218 | 0.096 |
| **+LF/HF** | 0.628 | 0.561 | 240.51 | 0.304 | 0.028* |
| **+LyapExp** | 0.621 | 0.553 | 241.14 | -0.297 | 0.037 * |
| **+ApEn** | 0.616 | 0.547 | 241.60 | -0.269 | 0.046* |
| **+Composite Mean-Z** | 0.675 | 0.618 | 235.84 | -0.390 | 0.003** |

*Note.* N=34. Each model was adjusted for baseline BPRS score, age, sex, and medication load. The clinical baseline model included only these covariates. Each subsequent model added a single measure separately. β values represent standardized regression coefficients for the added predictor. R<sup>2</sup> and adjusted R<sup>2</sup> refer to the full regression model. p values correspond to the added measure being tested with $p < 0.05^*$ , $p < 0.01^{**}$ .

